# Cohort profile of the ICMR-Stillbirth Pooled India Cohort (ICMR-SPIC): Estimating Prevalence, Analyzing Risk Factors, and Developing Prediction Models for Stillbirths in India

**DOI:** 10.1101/2025.01.16.25320656

**Authors:** ICMR-SPIC Consortium, Reema Mukherjee

## Abstract

**Purpose:** Stillbirth is a significant public health problem in India, yet comprehensive epidemiological data on its prevalence and risk factors are lacking. This initiative develops a pooled dataset from 10 well-characterized pregnancy cohorts across urban and rural India to estimate the national prevalence of stillbirths, identify risk factors and their population-attributable fractions, and develop a predictive risk stratification model for evidence-based clinical decision-making and interventions in high-risk pregnancies.

**Participants:** Pregnant women were recruited from the health facilities and community settings during the antenatal period. Recruitment spans four urban, four rural, and two mixed urban-rural sites, ensuring diversity in geographic and demographic representation.

**Findings to Date:** The ICMR stillbirth pooled India cohort (ICMR-SPIC) comprises 229,695 pregnant women. The mean (standard deviation) maternal Age at recruitment was 23.5(3.3) years. 30.5% were underweight (BMI <18.5 kg/m^2^) and 16.8% were overweight or obese (BMI ≥23 kg/m^2^). Short stature (<145 cm) was observed in 6.9% of participants. The mean (SD) gestational Age at birth was 38.7 (2.5) weeks. A third of the participants (33.3%) experienced moderate to severe anaemia during pregnancy (Hb<9.5g/dL), 52.8% were multiparous, and 22.5% conceived within 18 months of their previous childbirth. Core maternal risk factors such as short stature, BMI, parity, prior stillbirths, and anaemia during pregnancy were recorded in all cohorts. Additional variables, including gestational weight gain, preeclampsia/eclampsia, antepartum hemorrhage, and fetal distress, were available for over 80% of the cohorts, ensuring robust data coverage for risk factor analysis and modeling.

**Future Plans:** ICMR-SPIC will be used to conduct individual-level pooled data analyses to estimate prevalence, identify key risk factors, and develop predictive models for stillbirths. Findings will inform policies, clinical guidelines, and targeted interventions for high-risk pregnancies. The harmonized ICMR-SPIC dataset is a landmark collaborative effort to advance maternal and newborn health in India.

**STRENGTHS AND LIMITATIONS:**

- The harmonized **ICMR-SPIC pooled dataset** is the largest and most comprehensive resource for investigating the prevalence and determinants of stillbirths in India. It represents diverse geographical regions, encompassing both urban and rural recruitment sites, and provides a broad demographic spectrum.
- **Gestational Age at birth** was objectively determined using ultrasound measurements or last menstrual period data, enabling accurate categorization of stillbirths.
- The **prospective cohort design** facilitates identification of at-risk populations by providing demographic data across most cohorts and enabling longitudinal tracking of some modifiable risk factors.
- Despite rigorous efforts to harmonize data, variations in the measurement methods for certain modifiable risk factors (e.g., reproductive tract infections, preeclampsia, and haemoglobin concentrations) may result in residual misclassification, potentially affecting the precision of some risk analyses.
- A notable limitation of the dataset is the lack of detailed data in several cohorts to distinguish between **antepartum** and **intrapartum stillbirths**, or assess the quality of care during labour and childbirth. This restricts the ability to provide robust prevalence estimates or identify specific determinants for the two types of stillbirths.

## INTRODUCTION

The World Health Organization defines stillbirth as a baby born with no signs of life after 28 weeks of gestation or with a birthweight of less than 1000 grams[1]. Stillbirths before the onset of labor are classified as antepartum stillbirths, whereas those during labor and childbirth are grouped as intrapartum stillbirths. The Every Newborn Action Plan endorsed by the World Health Assembly in 2014 set a target of reducing the stillbirth rate to <12/1000 total births by 2030[1]. Furthermore, India pledged to reduce stillbirth and early neonatal mortality rates to <10/1000 births by 2030 through a focused strategy proposed in the 2014 India Newborn Action Plan[2]. Despite notable progress, most low- and middle-income countries (LMICs), including India, remain off track to achieve global targets for stillbirth reduction. The Global Burden of Disease study confirmed that India contributed the highest number (397,300) of stillbirths globally in 2021[3,4]. Over the past two decades, India has achieved an average annual reduction rate (ARR) of 4% in stillbirth rates, culminating in a 53% decline in2019 compared to 2000 (29.6 stillbirths per 1,000 total births in 2000 vs. 13.9 in 2019). However, the most recent estimates indicate that the burden remains unacceptably high, underscoring the need for intensified efforts to enhance maternal and perinatal healthcare systems to address persistent inequities [5,6].

Several challenges must be addressed to achieve the goal of reducing stillbirths in India. Notably, stillbirth targets are absent from global policy agendas, including the Sustainable Development Goals, and the definition of stillbirth varies across healthcare contexts, leading to misclassification and impeding international comparisons (Supplementary Table 1). This variation also contributes to discrepancies in stillbirth prevalence reported in different Indian registries[7,8]. Additionally, mechanisms for documenting stillbirths in LMICs, including India, remain suboptimal. For instance, the National Family Health Survey in India conflates stillbirths, miscarriages, and abortions as it relies on maternal self-reports[7], often introducing bias due to low maternal education and knowledge about stillbirths. Furthermore, there is limited epidemiological evidence on the burden and risk factors for stillbirths across India’s diverse regions[9,10], which is essential for designing tailored interventions. Generating comprehensive evidence on the prevalence and risk factors forstillbirths at a national scale necessitates coordinated, large-scale efforts that surpass the capacity of individual investigators, requiring multi-institutional collaboration, standardized methodologies, and robust data systems.

**Table 1.**
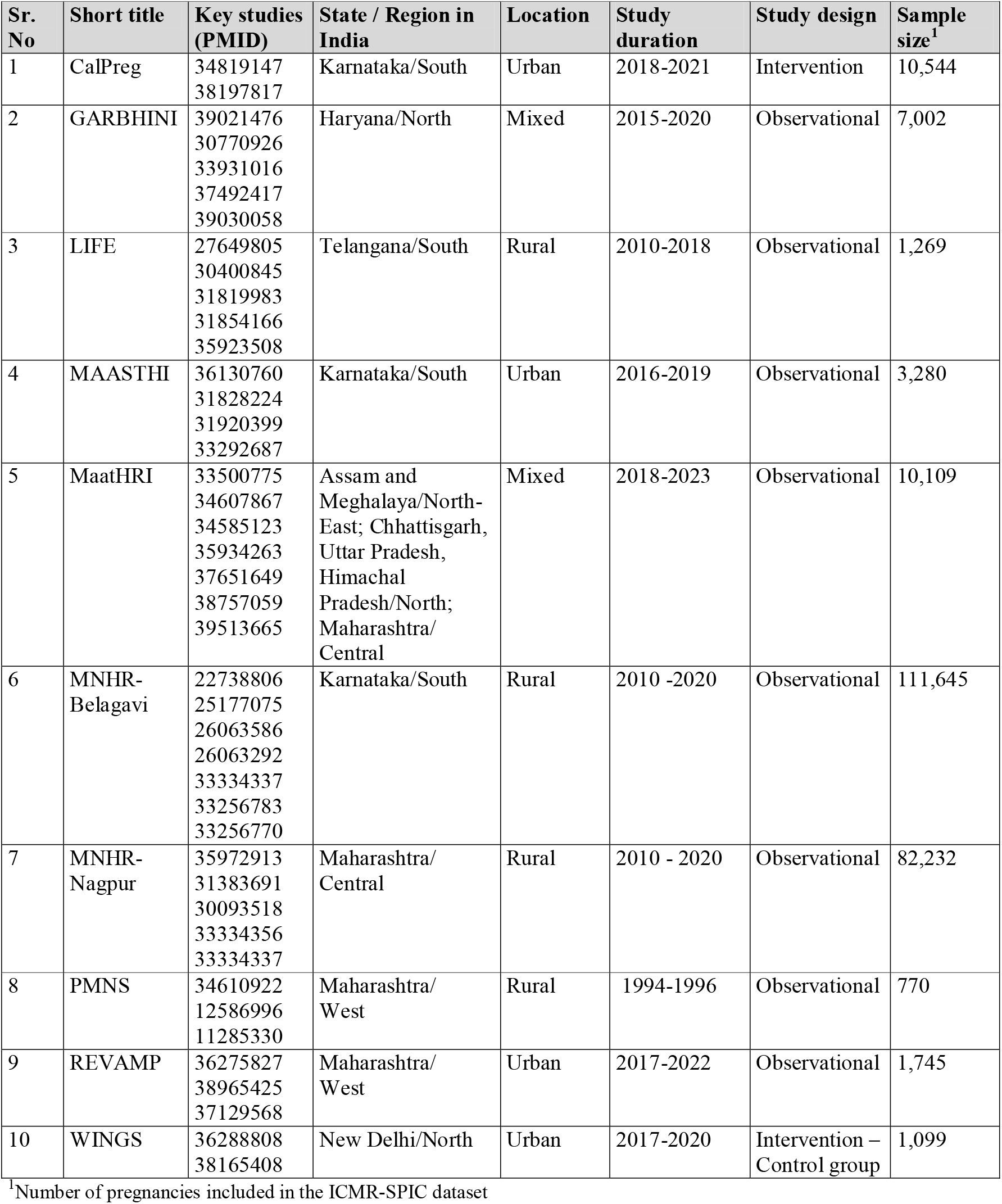
Details of pregnancy cohort studies included in the ICMR-SPIC harmonised dataset.

A collaborative, team-based approach is essential for generating robust evidence on stillbirths in India. The ICMR formed a consortium of pregnancy cohort studies to generate a harmonized dataset to estimate stillbirth prevalence, identify risk factors, and develop models to predict pregnancies at high risk of stillbirths in India. This initiative also aims to standardize the definition of stillbirth, enabling accurate burden estimation and advocacy. By aligning with India’s Every Newborn Action Plan, this effort is pivotal in developing evidence-based policies, interventions, and clinical guidelines to reduce preventable stillbirths.

Here, we outline the process of harmonizing data from multiple pregnancy cohorts to develop the Indian Council of Medical Research-Stillbirth Pooled India Cohort Dataset (ICMR-SPIC) and provide a concise description of the cohort profile. The ICMR-SPIC aims to estimate the national stillbirth rates in India, assess the associations between specific risk factors and stillbirths to evaluate their relevance for the Indian population and calculate the population attributable fraction for each risk factor to identify those with the most significant impact. Additionally, we will develop a risk prediction model for the early identification of pregnancies at high risk of stillbirth. This comprehensive approach provides a robust framework for generating actionable insights to reduce stillbirth rates in India.

## METHODS

### Selection of studies

In 2023, the ICMR coordinated forming of a consortium of investigators leading pregnancy cohorts in India, with the primary goal of pooling and harmonising data across all existing relevant cohorts. The dataset will be used for estimating the burden and determinants of stillbirths in India at the national level and for developing and validating a risk prediction model for identifying pregnancies at high risk of stillbirths that could benefit from targeted interventions. Ten investigator groups managing pregnancy cohorts joined the ICMR-SPIC consortium; details are provided in Supplementary Table 2. The consortium commenced its work in April 2024 after confirming the involvement of researchers, the availability of ethics and regulatory approvals, and signing agreements for sharing de-identified cohort data with the ICMR.

**Table 2.** List of variables and harmonised definitions.

| Sr. No. | Variable | Harmonised variable definition |
| --- | --- | --- |
| **Outcome measure** |  |  |
| 1 | Stillbirth | categorical; Yes/No/Not collected |
| **Maternal sociodemographic factors** |  |  |
| 1 | Maternal Age | continuous; completed years |
| 2 | Location | categorical; urban/rural/Not collected |
| 3 | Maternal education in years | continuous; completed years; level of maternal education (primary, secondary, etc, if completed years not available) |
| 5 | Consanguineous marriage | categorical; Yes/No/Not collected |
| 6 | Fuel type for cooking or heating | categorical; LPG/Natural gas/Kerosene/Coal/Charcoal/Wood/Dung cakes/Straw/Shrub/Grass/Agricultural crop waste/Biogas/Other/Combination of any of the above |
| 7 | Source of drinking water | categorical; Piped water into dwelling/ Public tap/Tubewell, borehole, or hand pump/Open well/Closed well/Tanker truck/Surface water/Bottled water/Rain water/ Other/ Combination of any of the above |
| **Maternal health history** |  |  |
| 1 | Weight (during pregnancy) | continuous; in kg |
| 2 | Height (during pregnancy) | continuous, in cm |
| 3 | Biceps skinfold thickness | continuous, in cm |
| 4 | Triceps skinfold thickness | continuous, in cm |
| 5 | Subscapular skinfold thickness | continuous, in cm |
| 6 | Mid-upper-arm circumference | continuous, in cm |
| 7 | Parity | discrete; count |
| 8 | Inter-pregnancy interval | continuous; completed months |
| 9 | Previous history of stillbirth | categorical; Yes/No/Not applicable/Not collected |
| 10 | History of previous abortion | categorical; Yes/No/Not applicable/Not collected |
| 11 | Prior caesarean section | categorical; Yes/No/Not applicable/Not collected |
| 12 | Pre-existing hypertension | categorical; Yes/No/Not collected |
| 13 | Family history of diabetes/hypertension/ cardiovascular disorders | categorical; Yes/No/Not collected |
| 14 | Artificial reproductive techniques | categorical; Yes/No/Not collected |
| 15 | Infertility treatment | categorical; Yes/No/Not collected |
| **Maternal health behaviours** |  |  |
| 1 | Tobacco consumption | categorical; Yes/No/Not collected |
| 2 | Alcohol consumption | categorical; Yes/No/Not collected |
| 3 | Passive smoking | categorical; Yes/No/Not collected |
| 4 | Number of ANC visits | discrete; count |
| **Maternal health during pregnancy** |  |  |
| 1 | Stress or depressive symptoms | continuous; raw score from tool of choice (EPDS/PHQ-9) |
| 2 | Overt diabetes | categorical; Yes/No/Not collected |
| 3 | Glycated haemoglobin (HbA1c) | continuous; % |
| 4 | Gestational diabetes (GDM) | categorical; Yes/No/Not collected |
| 5 | GDM time of diagnosis | continuous; completed weeks of gestation |
| 6 | Haemoglobin | continuous; g/dL |
| 7 | Haemoglobin – method of assessment | categorical; Autoanalyser/Point of care testing/Sahli's method/Photometric method/Not collected |
| 8 | Fasting plasma glucose | continuous; mg/dL |
| 9 | 1-hr plasma glucose: Oral glucose tolerance test | continuous; mg/dL |
| 10 | 2-hr plasma glucose: Oral glucose tolerance test | continuous; mg/dL |
| 11 | Systolic blood pressure | continuous; mmHg |
| 12 | Diastolic blood pressure | continuous; mmHg |
| 13 | Urinary protein | continuous; g/dL |
| 15 | Gestational hypertension (GH) | categorical; Yes/No/Not collected |
| 16 | Time of diagnosis: GH | continuous; completed weeks of gestation |
| 17 | Preeclampsia | categorical; Yes/No/Not collected |
| 18 | Time of diagnosis: Preeclampsia | continuous; completed weeks of gestation |
| 19 | Eclampsia | categorical; Yes/No/Not collected |
| 20 | Time of diagnosis: Eclampsia | continuous; completed weeks |
| 21 | Thyroid disorder | categorical; Hypothyroidism/Hyperthyroidism/Euthyroid/Not collected |
| **Maternal infections during pregnancy** |  |  |
| 1 | Asymptomatic bacteriuria | categorical; Yes/No/Not collected |
| 2 | Reproductive tract infection | categorical; Yes/No/Not collected |
| 3 | Syphilis | categorical; Yes/No/Not collected |
| 4 | Human immunodeficiency virus | categorical; Yes/No/Not collected |
| 5 | Malaria | categorical; Yes/No/Not collected |
| 6 | Rubella | categorical; Yes/No/Not collected |
| 7 | Varicella | categorical; Yes/No/Not collected |
| 8 | Toxoplasma | categorical; Yes/No/Not collected |
| 9 | Hepatitis B | categorical; Yes/No/Not collected |
| 10 | Tuberculosis | categorical; Yes/No/Not collected |
| 11 | Cytomegalovirus | categorical; Yes/No/Not collected |
| **Obstetric factors at the time of childbirth** |  |  |
| 1 | Date of childbirth | date variable; DD/MM/YYYY |
| 2 | Mode of childbirth | categorical : vaginal/assisted/caesarean |
| 3 | Place of childbirth | categorical; Institutional/Non-institutional/Not collected |
| 4 | Gestational Age (GA) at childbirth | continuous, in **weeks** |
| 5 | GA at childbirth in days | continuous, in **days** |
| 6 | Method of dating GA | categorical; Ultrasound sonography (USG)/Last menstrual period/Others |
| 7 | USG method of dating GA | categorical; Crown-Rump length/Other fetal biometry/Not applicable |
| 8 | GA at dating (weeks) | continuous, in weeks |
| 9 | GA at dating (days) | continuous, in days |
| 10 | Date of dating | date variable; DD/MM/YYYY |
| 11 | Ultrasound evidence of fetal heart activity just before onset of labour or rupture of membranes | categorical; Yes/No/Not collected |
| 12 | Perception of fetal movements just before onset of labour or rupture of membranes | categorical; Yes/No/Not collected |
| 13 | Obstructed or prolonged labour or failure to progress | categorical; Yes/No/Not collected |
| 14 | Mal-presentation at childbirth | categorical; Yes/No/Not collected |
| 15 | Antepartum haemorrhage | categorical; Yes/No/Not collected |
| 17 | Amniotic fluid disorders (AFD) | categorical; |
|  |  | Oligohydramnios/Polyhydramnios/Normal/Not collected |
| 18 | Time of diagnosis: AFD | continuous; completed weeks of gestation |
| **Maternal blood biomarkers during pregnancy (available in a subset)** |  |  |
| 1 | Vitamin B12 | continuous; pg/mL |
| 2 | Folate | continuous;ng/mL |
| 3 | Ferritin | continuous;ng/mL |
| 5 | Soluble transferrin receptor | continuous;mg/mL |
| 6 | Vitamin D | continuous;ng/mL |
| 7 | Vitamin B6 | continuous;ng/mL |
| 8 | Zinc | continuous;ug/dL |
| 9 | Selenium | continuous;ug/L |
| 10 | C-reactive protein (CRP) | continuous;mg/L |
| 11 | hs-CRP | continuous;mg/L |
| 12 | Calcium | continuous;mg/dL |
| 13 | Magnesium | continuous;mg/dL |
| 14 | Methyl malonic acid | continuous;ng/mL |
| 15 | Cortisol | continuous;μg/dL |
| 16 | Total cholesterol | continuous;mg/dL |
| 17 | Low-density lipoprotein (LDL) | continuous;mg/dL |
| 18 | High-density lipoprotein (HDL) | continuous;mg/dL |
| 19 | Triglycerides (TGL) | continuous;mg/dL |
| 20 | Homocysteine | continuous;μmol/L |

For a cohort study to be included in the pooled analysis, the study should have fulfilled the following criteria:

1. The study must be conducted in either urban or rural India. Pregnant women could be recruited in health facilities or community settings.
2. The cohort studies should have appropriate ethics and regulatory approvals, including participant consent for sharing data with third parties for secondary analysis.
3. Pregnant women should have been recruited before the birth of their child and followed longitudinally until a birth outcome (live birth, stillbirth, medical or spontaneous abortion) was recorded.
4. The cohort studies must provide detailed descriptions of the study methods, including recruitment and sampling strategies, inclusion and exclusion criteria, and detailed definitions for all the variables shared with the consortium, to enable data harmonisation and accurate interpretation of the results.
5. The dataset must include a core set of “required” variables, such as gestational Age at childbirth (GA), along with methods determining GA (Last Menstrual Period or ultrasonography), and a set of sociodemographic variables, such as maternal Age at childbirth, education levels, etc.
6. While desirable, the availability of details of medical conditions, obstetric complications, and behavioral factors were not considered mandatory.

### Data sources for ICMR-SPIC

Ten datasets were included in the pooled ICMR-SPIC database (see Table 1 for details), spanning17 sites across nine Indian states representing North, West, Central, South, and North-Eastern India. One study (MaatHRI) included study sites in the North, North-East, and Central Indian regions. Eight out of ten studies followed an observational study design. Among the two intervention studies that were included, the WINGS cohort contributed data only from the control arm. In contrast, the CalPreg cohort contributed data from both the control and intervention arms, as the intervention differed only in the dose of calcium administered during pregnancy. Furthermore, four out of ten cohorts recruited participants from community settings and the remaining six were hospital-based . While there was an equal representation of the *number of sites* contributing data from urban and rural areas (four each from urban and rural areas, and two with mixed urban and rural populations, see Table 1), the majority of participants, *in absolute numbers* (91.8%, Table 3), were from rural areas.

**Table 3.**
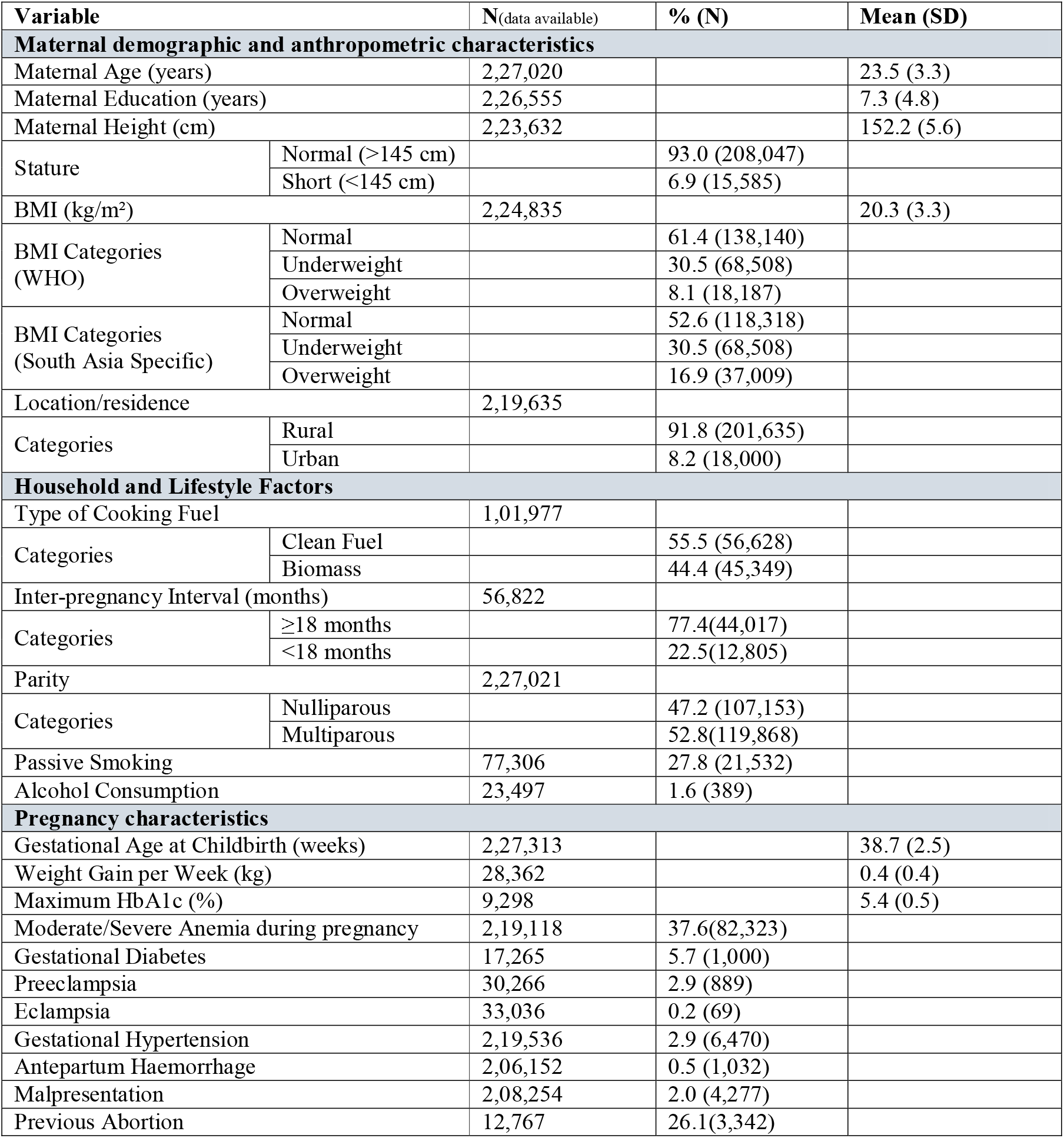
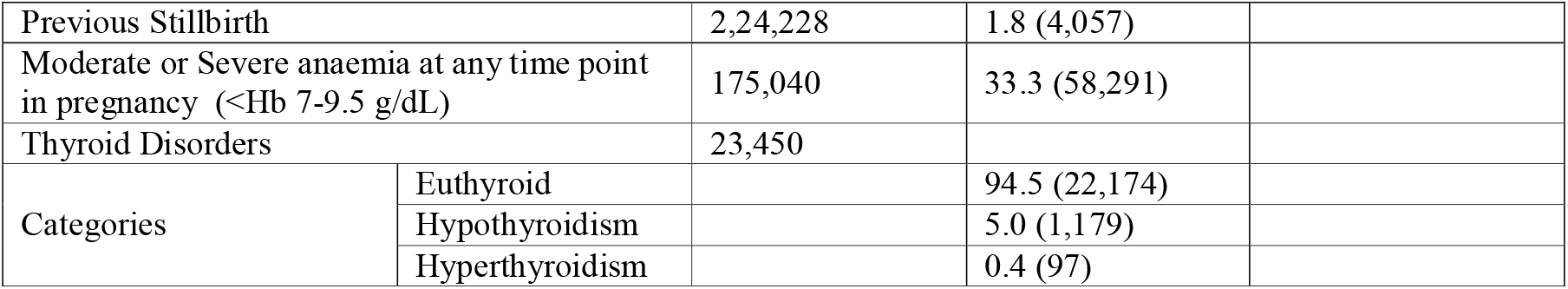
Sociodemographic and health characteristics of the ICMR-SPIC Cohort.

### Data harmonization and data cleaning

A preliminary draft of the data dictionary and codebook that listed the essential and desirable variables to be included in the ICMR-SPIC dataset was prepared including their tentative definitions and harmonised variable names. This draft was shared with all the consortium partners for review and comments. The consortium members discussed, modified, and mutually agreed on the final list of variables and their definitions. The updated data dictionary and codebook was shared with all the consortium members for mapping and recoding their data into the final data template. Each cohort performed thorough data cleaning to adhere to the definitions and codes, and prepared detailed notes on how data on each variable was collected and categorized to minimise variations in measurement methods that could impact the interpretation of results. If the teams were unable to adhere to the harmonised definitions, they provided a detailed description of the discrepancy compared with the requested format, definition, or assessment method. Each cohort then uploaded the cleaned and final dataset along with the annotated codebook on a secure webserver managed by ICMR.

The core statistical analysis team then reviewed each dataset to ensure fidelity to the harmonization template. Data managers of the respective cohorts corrected any errors highlighted by the statistical team. Some cohorts had recruited the same participant across multiple pregnancies. In these cases, a decision was made to represent each row as a unique pregnancy, with one column linking participants across pregnancies whenever possible. Multifetal gestations were reported as individual rows for each fetus, with a column indicating singleton or multiple gestation for each observation. This allowed the team to assess the outcome of each birth from all pregnancies while accounting for the fact that not all participants were independent in the combined dataset, which will be accounted for using appropriate statistical methods during the analysis. Summary statistics and histograms/bar graphs were plotted for each derived variable and their component raw variables to identify outliers and missing data were examined. Codes were written to check for range and logical errors for each variable, and any data or coding errors were corrected with help from the respective cohort’s data managers. The final verified datasets were then sequentially appended to the master dataset one at a time using code prepared in STATA version 16 (Stata Corp., College Station, USA) or R version 3.3.3 (R Core Team, 2023; https://www.R-project.org/).The only major exclusion was missing outcome data (live/stillbirth) or if the fetus was naturally or medically aborted. The harmonized dataset was accessible to all consortium members through password protected access controls to the secure server.

### Definitions

The final list of variables and their definitions are presented in Table 2. Gestational Age (GA) calculation was prioritised from ultrasound data when available and from last menstrual period if ultrasound data was not available. GA was used to determine what proportions of births were preterm and to calculate the time-period (or gestational Age) of measurement of each risk factor during the pregnancy. This was done to enable adjustments for time-varying exposures in statistical models, as many risk factors are known to have differential effects on stillbirth outcome depending on the stage at which they affect pregnancy.

#### Primary outcome measure: Stillbirth

The team made a decision to adopt the WHO definition of stillbirth (birth of a fetus without any sign of life at or after 28 weeks of gestation), which is recommended for international comparisons.

### Principles and plans for statistical analysis

A detailed study flow diagram outlining the analytical decisions that progressed from the total participant pool across all cohorts to the final analytic dataset is presented in Supplementary Figure 1. For each objective, a comprehensive statistical analysis and reporting plan was formulated in collaboration with the Technical Advisory Group of the ICMR-SPIC consortium. This plan delineated the statistical techniques, underlying assumptions, and procedural steps, ensuring systematic and transparent analyses (details will be reported in subsequent papers). The harmonized dataset will be analyzed using two complementary approaches to generate the most robust evidence: (1) a one-stage meta-analysis, an individual-level pooled analysis of all available data, and (2) a two-stage meta-analysis: a meta-analysis of cohort-specific summary data to examine and account for between-study heterogeneity among the cohorts.

Using a weighted sample, the stillbirth rate (SBR) will be calculated as the number of stillbirths divided by the total number of births expressed per 1,000 total births. To account for differences in sample sizes across cohorts, each cohort will be weighted, with weights computed as the inverse of the ratio of the individual cohort sample size to the overall pooled cohort sample size. The total number of births will be defined as the sum of live births (regardless of gestational Age) and stillbirths. SBR at the national level will be reported along with the 95% confidence intervals.

#### Assessment of Risk factors for stillbirths in India

To assess the association and estimate the risk ratios of various sociodemographicand antenatal risk factors for stillbirth, a modified mixed-effects model will be utilised with each cohort included as a random effect. Directed acyclic graphs (DAGs) will be drawn to understand the pathways of the known risk factors. To quantify the impact of each risk factor, the population attributable fraction (PAF) will be estimated using Miettinen’s formula[11], expressed as: *PAF* = *P*_*e*_ (*RR - 1*)/*RR*, where *P*_*e*_ is the proportion of stillbirth cases exposed to the risk factor, and RR is the adjusted relative risk for that risk factor.

#### Development and internal validation of a risk prediction model to identify pregnancies at high risk of stillbirth

A clinical prediction model will be developed to predict the risk of stillbirth in pregnant women visiting healthcare facilities using their baseline (fixed) and modifiable risk factors, aimed to support clinicians in medical decision-making. The optimal set of predictors that contribute significantly to predicting stillbirthswill be identified by domain knowledge-based and data-driven approaches.

Using a naive Bayesian framework, a dynamic model [12] will be used to dynamically assess the personalised risk of stillbirth. The initial baseline probability will be derived from the estimated prevalence of stillbirth for the study population. Thereafter, conditional probabilities will be computed for each new predictor using Bayes Theorem to update the risk of stillbirth for each pregnant woman. The model will be internally validated on the ‘ left-out’ dataset. The model will be evaluated for quantifying the error in prediction (root mean squared error, mean absolute error, and calibration-in-the-large (CITL), discrimination ability using the area under the receiver operating characteristics curve and decision curve analysis.

## PATIENT AND PUBLIC INVOLVEMENT STATEMENT

No patients or members of the public were directly involved in the design, conduct, or analysis of this secondary data analysis. However, the ICMR-SPIC consortium includes representatives from India’s research, practice, and policy communities to ensure that the study aligns with national health priorities and addresses key public health concerns. Dissemination efforts will focus on engaging future mothers and their families, the general public, non-governmental organizations dedicated to preventing stillbirths and improving maternal and child health, and other relevant stakeholders. Study findings will be communicated through diverse channels, including local audio-visual media, print media, and social media platforms, with messages specifically tailored to inform future mothers and their families about stillbirth risk factors and effective strategies for prevention and management.

## CHARACTERISTICS OF THE COHORT PARTICIPANTS

The harmonized ICMR-SPIC dataset comprises individual-level data from a large sample of 2,29,695pregnant womenon maternal sociodemographic, health, lifestyle, and household factors, as well as characteristics of previous and current pregnancies and objective measures of birth outcomes (Table 3).

### a) Maternal sociodemographic and anthropometric characteristics

The mean (standard deviation or SD) maternal Age at enrolment was 23.5(3.3) years. Education duration varied widely, with a mean (SD) of 7.3 (4.8) years, which is expected for the profile of women visiting public health facilities in India or residing in urban-poor or rural community settings. The mean(SD) maternal height was 152.2(5.6) cm, with 6.9% being of short stature (<145 cm), a known risk factor for adverse pregnancy outcomes, including stillbirth[13]. Using criteria specified for the South Asian population, 30.5% of women were underweight (< 18.5 kg/m^2^) and 16.8% were overweight (≥ 23 kg/m^2^), highlighting the double burden of malnutrition in India - persistent issues of undernutrition along with an emergent problem of overweight and obesity. The majority of the participants (91.8%) live in rural areas.

### b) Maternal household and lifestyle factors

Nearly half the cohort (44.4%) used biomass fuels, known to contribute to indoor air pollution and respiratory health issues.22.5% of mothers had an inter-pregnancy interval of <18 months, which has been suggested to be associated with a higher risk of adverse maternal and child health outcomes. However, data was available only for a quarter of the total sample for this variable (N=56,822). The distribution of parity was balanced, with 47.2% of women being nulliparous and 52.8% multiparous, the former being reported as a risk factor for stillbirths[14]. 27.8% and 1.6% reported being exposed to passive smoking and consuming alcohol during pregnancy, respectively. However, data for these factors were available in a small subset of the total population (Table 3).

### c) Pregnancy characteristics

Although information about the history of abortion was available only for 12,767 women, 26.1% of these women reported having experienced a previous abortion. In contrast, information about previous stillbirths was available for almost all participants (N=2,24,228), with 1.8% reported experiencing at least one previous stillbirth. The mean (SD) gestational Age at childbirth was 38.7 (2.5) weeks, and gestational weight gain per weekwas0.4 (0.4) kg (N=28,362).The prevalence of pregnancy complications computed from smaller subsets of the data was noted as follows: gestational diabetes (5.7%, N=17,265), gestational hypertension (2.9%, N=2,19,536), preeclampsia (2.9%, N=30,266), eclampsia (0.2%, N=33,036), antepartum hemorrhage (0.5%, N=2,06,152), and fetal malpresentation (2.0%, N=2,08,254).The data also revealed substantial and concerning rates of anemia, with 33.3% of women having moderate or severe anemia during pregnancy. Additionally, the data indicated that 5% of women had thyroid disorders (diagnosed with hypothyroidism).

## DISCUSSION

The ICMR-SPIC dataset represents a landmark collaborative initiative consolidating data from 10 well-characterized pregnancy cohorts spanning diverse regions of India. This harmonized dataset, encompassing 229,695 participants from both urban and rural settings, is the largest of its kind in the country, which will facilitate robust analyses of stillbirth prevalence, associated risk factors, and predictive models for high-risk pregnancies. Using standardized methodologies and harmonized definitions enhances the dataset’s reliability, allowing national and regional estimates to support evidence-based policy formulation and targeted intervention design. The findings will underscore critical maternal health challenges leading to a high burden of stillbirth in the country, including a high prevalence of malnutrition, anemia, pregnancy complications such as gestational diabetes and preeclampsia, and significant exposure to passive smoking and biomass fuels. However, the dataset highlights notable gaps in representation, mainly from eastern India and tribal populations, and inconsistencies in data collection methods across cohorts. These factors necessitate careful consideration during data analysis and interpretation to ensure accurate and meaningful insights. Despite limitations, including incomplete data on pre-conception factors, intrapartum care, and variability in measurement methods, the scale and scope of the ICMR-SPIC dataset offer an unprecedented opportunity to develop predictive models and design context-specific interventions and is a major step forward in collaborative research within India.

## CONCLUSION

The ICMR-SPIC demonstrates the value of large-scale collaborative data harmonization approaches to address critical public health challenges like stillbirth in India. By pooling data from diverse pregnancy cohorts, this unique effort enables robust, generalizable insights into the prevalence and region-specific risk factors for stillbirths, and facilitates the development of a prediction model to identify pregnancies at high risk of stillbirths. These efforts will inform evidence-based clinical guidelines, interventions, and policymaking, thereby addressing the goal of reducing rates of preventable stillbirths in India, and achieving the national and global targets.

## FURTHER DETAILS

### Data availability statement

All contributing research teams have acknowledged that the pooled data can only be used for collaborative activities within the ICMR-SPIC consortium, with no transfer of ownership.

### Funding

This study was funded by the Indian Council of Medical Research to create a harmonised pooled dataset across ten pregnancy cohorts in India and to conduct secondary analysis using this dataset.

### Contributors

ICMR conceptualized the study. RC, RT, and RM designed the study with input from all members of the ICMR-SPIC consortium and the Technical Advisory Group (TAG). RM, AY, and JM were responsible for data cleaning, data preparation, and statistical analyses, supervised by RC, RT and TAG. All authors contributed to data acquisition and interpretation of results. DM and RC drafted, and MN edited the manuscript; all authors reviewed it and provided consent for publication of the final version.

### ICMR-SPIC Consortium

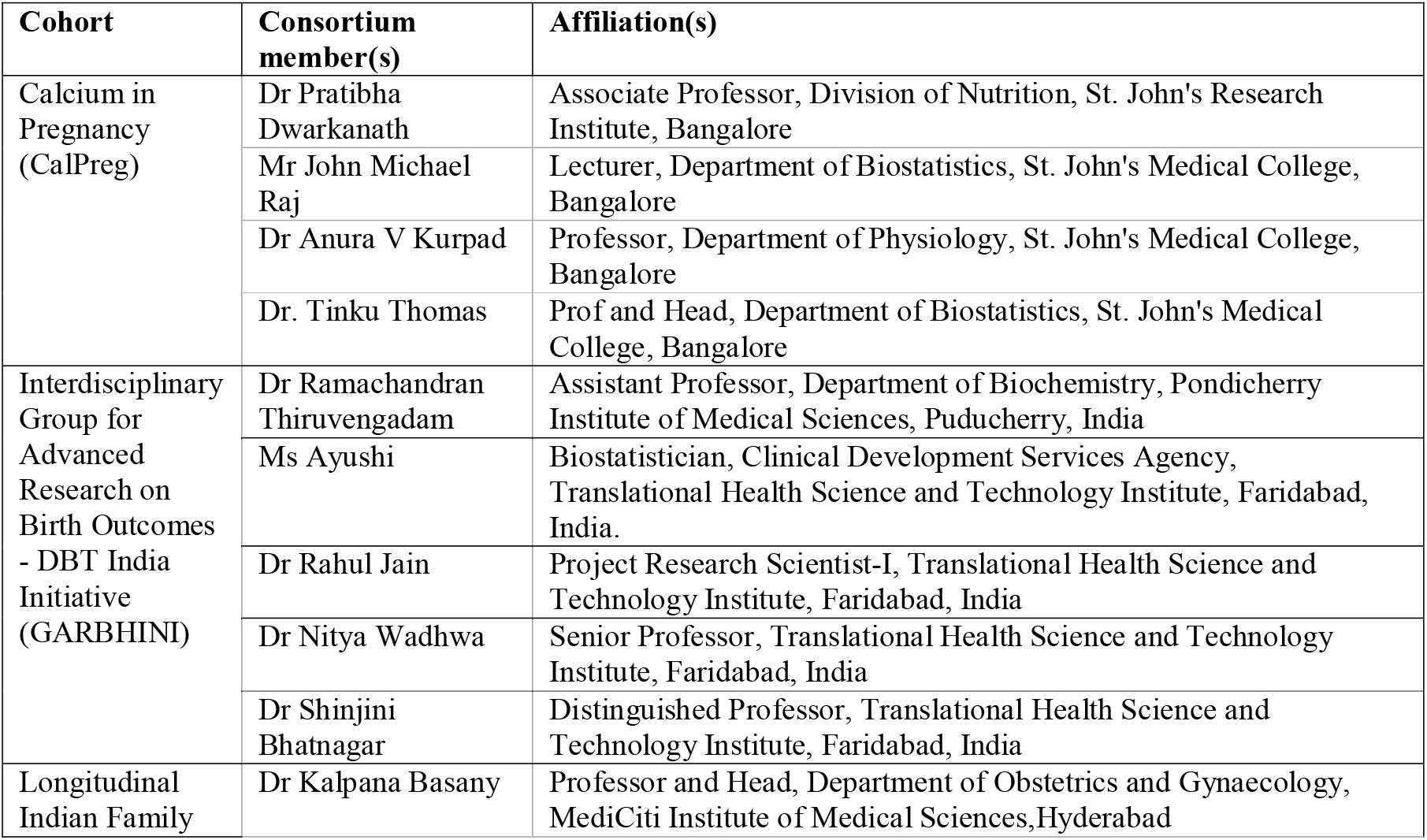
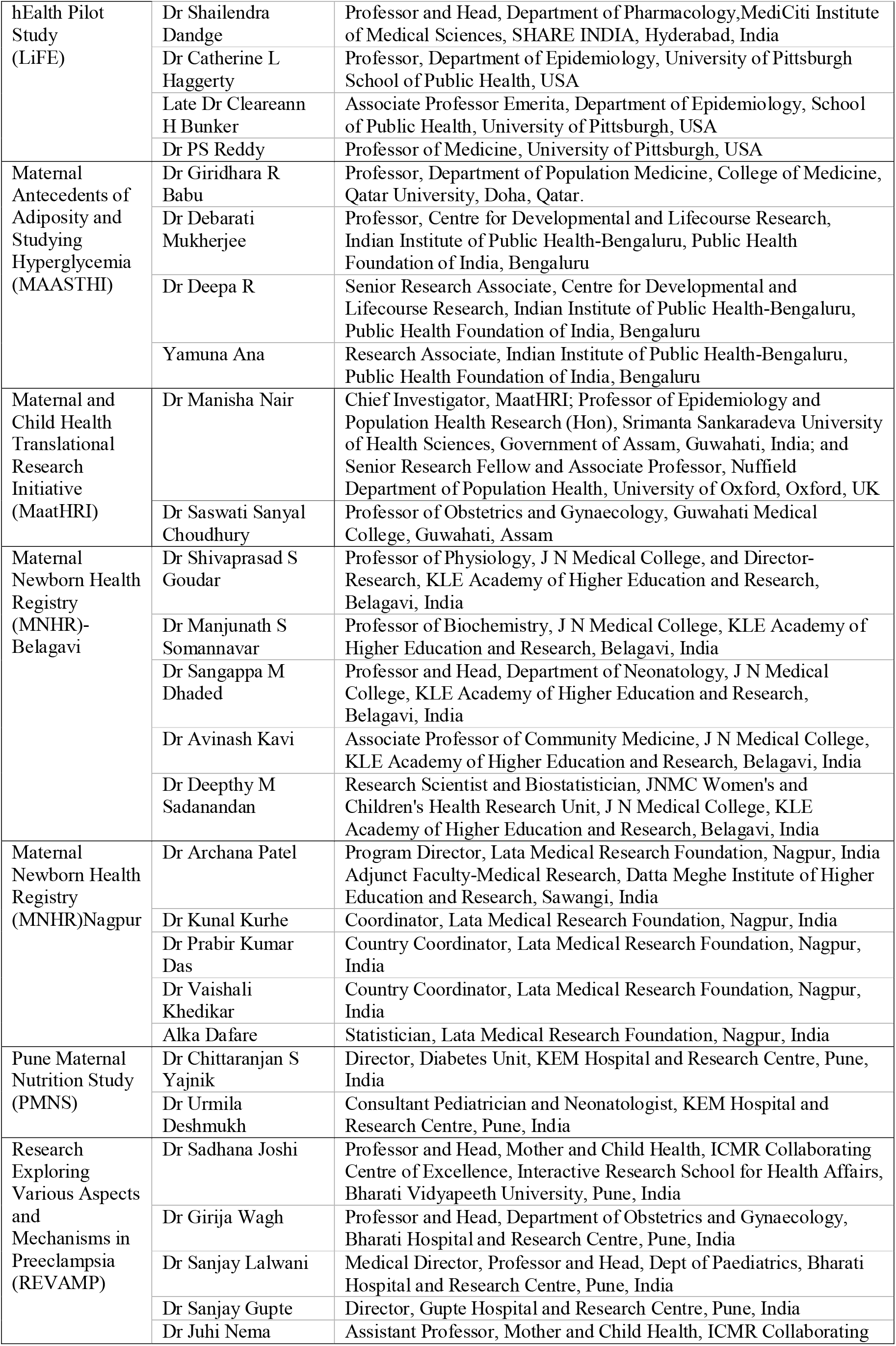
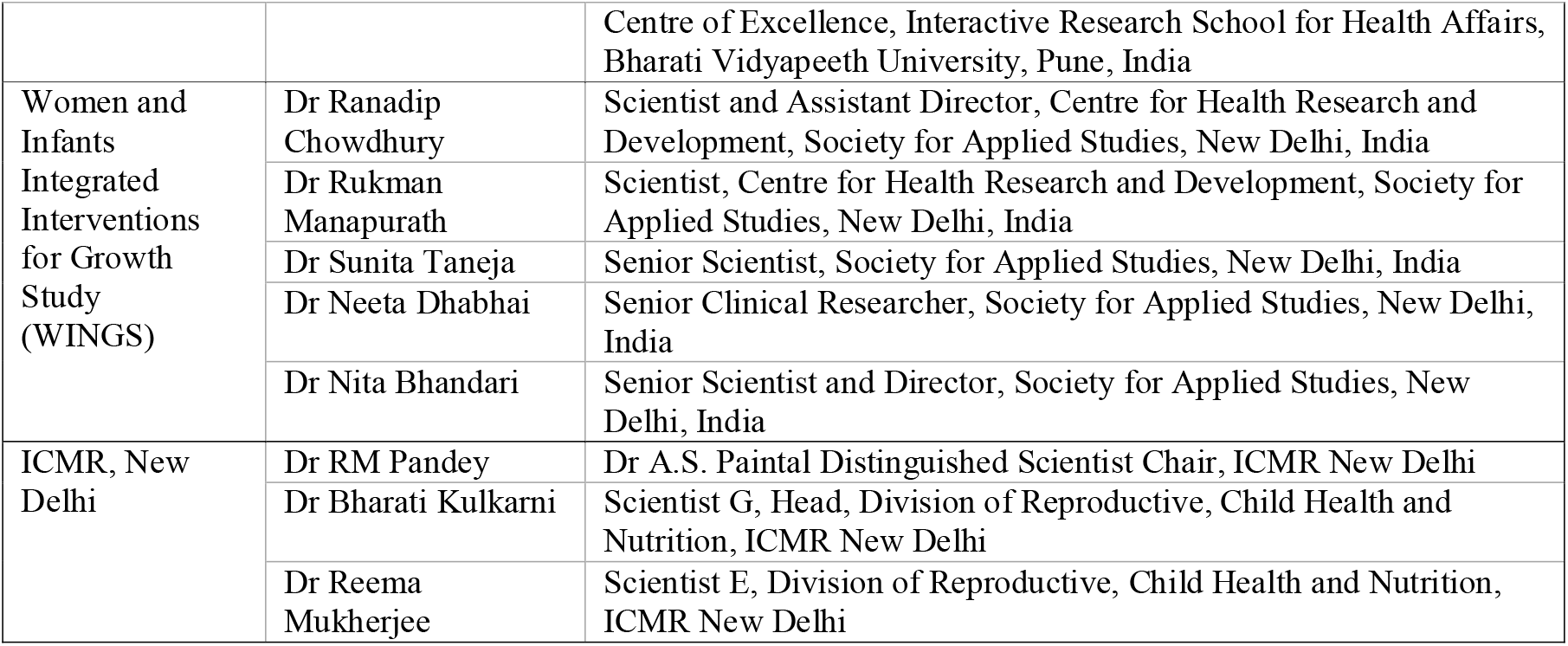

## Acknowledgement

We sincerely thank the study participants for sharing valuable information and biological samples for the conduct of this study. Additionally, each cohort team thanks the following research staff for supporting the preparation of the harmonized dataset. We are thankful to the primary funding agencies of the cohorts for granting permission to share the data for this exercise.

The ICMR-SPIC consortium would also like to thank the technical advisory group members for their support in conceptualizing the specific objectives and statistical analysis plan for this study.

| Cohort(in alphabetical order) | Acknowledgement |
| --- | --- |
| CalPreg | Dr. Wafaie W. Fawzi, Department of Global Health and Population, Harvard T.H. Chan School of Public Health, Boston, Massachusetts, USA (the primary recipient of the multicentric grant), for allowing the use of the India trial data for this analysis. |
| GARBHINI | Department of Biotechnology, Government of India for permitting to participate in the consortium GARBH-Ini study team (Shinjini Bhatnagar, Nitya Wadhwa, Uma Chandra MouliNatchu, Bhabatosh Das, Pallavi S Kshetrapal, Shailaja Sopory, Ramachandran Thiruvengadam, Sumit Misra, Dharmendra Sharma, Kanika Sachdeva, Amanpreet Singh, Balakrish G Nair, Satyajit Rath, and Vineeta Bal (Translational Health Science and Technology Institute, Faridabad, Haryana, India); Alka Sharma, Sunita Sharma, Umesh Mehta, and Brahmdeep Sindhu (Gurugram Civil Hospital, Gurugram, India); Pratima Mittal, Rekha Bharti, Harish Chellani, Rani Gera, Jyotsna Suri, Pradeep Debata, and Sugandha Arya (Vardhman Mahavir Medical College & Safdarjung Hospital, New Delhi, India); Arindam Maitra (National Institute of Biomedical Genomics, Kalyani, India); Tushar K Maiti (Regional Centre for Biotechnology, Faridabad, India); Dinakar M Salunke (International Centre for Genetic Engineering and Biotechnology, New Delhi, India); Nikhil Tandon, Yashdeep Gupta, Alpesh Goyal, Smriti Hari, Aparna Sharma K, and Anubhuti Rana (All India Institute of Medical Sciences, New Delhi, India); SiddarthRamji and Anju Garg (Maulana Azad Medical College, New Delhi, India); Ashok Khurana (The Ultrasound Lab, Defence Colony, New Delhi, India); Reva Tripathi (Sitaram Bhartia Institute of Science and Research, New Delhi, India); Rakesh Gupta and Partha P Majumder (Government of India); Himanshu Sinha and Raghunathan Rengasamy (Indian Institute of Technology Madras, Chennai, India); Vineeta Bal (Indian Institute of Science Education and Research, Pune, India); Pratima Mittal (Amrita Institute of Medical Sciences & Research Centre, Faridabad, India); Uma Chandra MouliNatchu and Harish Chellani (Center for Health Research and Development, Society for Applied Studies, New Delhi, India); and Ramachandran Thiruvengadam (Pondicherry Institute of Medical Sciences, Puducherry, India).Members listed in the supplementary)<br>Mr Ashu Sharma, Technical officer-II, Translational Health Science and Technology Institute, Faridabad, India, |
| LiFE | Mr Purushotham Reddy R, Head, Information Technology, Mr Narender Konga, Data Assistant, and Mrs Deepa V Project Manager, MediCiti Institute of Medical Sciences, SHARE INDIA, Hyderabad, India |
| MAASTHI | Ms Suganya S, Indian Institute of Public Health-Bengaluru, PHFI |
| MaatHRI | Dr Tuck Seng Cheng, University of Oxford; Ms RupanjaliDeka, Srimanta Sankaradeva University of Health Sciences, Guwahati, and all MaatHRI site collaborators, namely;Farzana Zahir, Carolin |
|  | Solomi V, Ashok Verma, Sereesha Rao, SaswatiSanyal Choudhury, GitanjaliDeka, Pranabika Mahanta, Swapna Kakoty, Robin Medhi, Shakuntala Chhabra, Anjali Rani, Amrit Bora, Indrani Roy, Bina Minzand, Omesh Kumar Bharti. Members of MaatHRI study team: Manisha Nair, Farzana Zahir, Carolin Solomi V, Ashok Verma, Sereesha Rao, SaswatiSanyal Choudhury, Gitanjali Deka, Pranabika Mahanta, Swapna Kakoty, Robin Medhi, Shakuntala Chhabra, Anjali Rani, Amrit Bora, Indrani Roy, Bina Minz, Omesh Kumar Bharti, Rupanjali Deka, Tuck Seng Cheng. |
| MNHR-Belagavi | Dr N V Honnunar, Field Research Officer, JNMC Women's and Children's Health Research Unit; Dr Richard J Derman, Associate Provost Global Affairs, Director Global Health Research, Professor OBGYN, Thomas Jefferson University, PA USA; Dr. Elizabeth M McClure and Ms. Janet Moore, RTI International, Durham, NC, USA ;Dr. Marion Koso-Thomas, *Eunice Kennedy Shriver* National Institute of Child Health and Human Development, Bethesda, MD, USA; Dr Ashalata Mallapur and Dr Umesh Ramadurg, S N Medical College, Bagalkot; Medical Officers of Department of Health and Family Welfare, Government of Karnataka: Dr B D Yalgudri, Dr S M Vijapur, Dr Shivanand S Mulakuri, Dr Vrushbendra D Hiremath, Dr Ramkrishna Yadgude, Dr M J Kumbar, Dr B N Kirankumar, Dr Salim Mujawar, Dr S G Shetter, Dr S C Mastiholi, Dr Udaya Kudachi, Dr Shaheed S Gadekai, Dr S M Chavadappanavar, Dr R G Patil, Dr Harsha Patil, Dr S G Kamble, Dr Prashant Gaikwad, Dr R A Makhandar, Dr S S Doddmani, Dr BapusahebPatil, Dr C S Patil, Dr Sukanya Handage, Dr R S Balikai, Dr Rajashekar Mudhkamath, Dr S SSiddannavar, Dr Anil S Salagure, Dr Rajendra K Kilabanur, Dr Arun V Halagatti, Dr Poornima Tallur, Dr Shridhar Pattar, Dr (Mrs) Kusuma Magi, Dr Vishwanath M Pattar, Dr Vikas B Parvaikar, Dr Bhagya Kshatri, Dr Ashok Kuntoji; Mr. Nagaraj Khade, Mr. Sachin Mastiholi, Mr. Vinayak Mhetri, Mr. Dayanand Shiroor, Mr. Kadappa Beniwadi |
| MNHR Nagpur | Dr. Patricia Hibberd, Department of Global Health, Boston University School of Public Health, Boston, MA, USA; Dr. Elizabeth M McClure and Ms. Janet Moore, RTI International, Durham, NC, USA; Dr. Marion Koso-Thomas, *Eunice Kennedy Shriver* National Institute of Child Health and Human Development, Bethesda, MD, USA; Officials, Department of Health and Family Welfare, Government of Maharashtra, India; Ms. Smita Puppallwar, Dr. Rakesh Kukde, Dr. Damodar Saliwkar, Dr. Shashank Parankar, Mr. Nitesh Meshram, Lata Medical Research Foundation, Nagpur, India. |
| PMNS | Dr Prachi Phadke and Onkar Deshmukh, KEM Hospital and Research Centre, Pune, India |
| REVAMP | Karuna Randhir, Interactive Research School for Health Affairs, Bharat Vidyapeeth University |
| WINGS | Kiran Bhatia, Society for Applied Studies, New Delhi, India |

## Technical Advisory Group of the ICMR-SPIC consortium

| Sr. No | Member name | Institution |
| --- | --- | --- |
| 1 | Dr RM Pandey (Chairperson) | Dr A.S. Paintal Distinguished Scientist Chair, ICMR New Delhi |
| 2 | Dr Anamika Gambhir | Department of Biotechnology, Govt. of India |
| 3 | Dr Shobhna Das | Ministry of Health and Family Welfare, Govt. of India |
| 4 | Dr Rakhi Dandona | Public Health Foundation of India, New Delhi, India |
| 5 | Dr Pranay Goel | Indian Institute of Science Education and Research, Pune, India |
| 6 | Dr Rajiv Sarkar | Indian Institute of Public Health, Shillong, India |
| 7 | Col (Dr) Arun Kumar Yadav | Office of Director General of Armed Forces Medical Services in India |
| 8 | Dr K Aparna Sharma | All India Institute of Medical Sciences, New Delhi, India |

## Disclaimer

The authors alone are responsible for the views expressed in this paper, and they do not necessarily represent the views, decisions, or policies of the institutions with which they are affiliated.

