## Supplemental Information for "Cohort profile of the ICMR-Stillbirth Pooled India Cohort (ICMR-SPIC): Estimating Prevalence, Analyzing Risk Factors, and Developing Prediction Models for Stillbirths in India"

**Supplementary Figure1:**Study flow depicting cohort and participant details in the final analytical dataset

Contacted for Data Sharing 13 teams

Signed Data Sharing Agreement 10 cohort teams

Received Data for Pooling: 10 teams 247,050 data points

Pooled Dataset: 10 teams 247,050 data points

Analysis Pooled Dataset 229,695 (93%)

Missing outcome data + abortions= 17,355 (7%)

3 excluded Administrative issues with data sharing agreement

**Supplementary Table 1:**Stillbirth(birth of a baby with no signs of life)as defined by key health organizations

| **Sr. No** | **Source** | **Gestational age at delivery** | **Other Criteria** | **Further classification** |
| --- | --- | --- | --- | --- |
| 1 | World Health Organization | ≥28 weeks | Birthweight: ≥1000 gms | Early stillbirth: 22-27 weeks  Late stillbirth: ≥28 weeks |
| 2 | Centre for Disease Control - USA | ≥20 weeks | Birthweight: ≥350gms |  |
| 3 | American College of Obstetricians and Gynecologists (ACOG) | ≥20 weeks | Birthweight: ≥ 500gms |  |
| 4 | National Health Service (NHS) - United Kingdom | ≥24 weeks |  | Late miscarriage:<24 weeks |
| 5 | International Classification of Diseases (ICD-10) | ≥22 weeks | Birthweight: ≥ 500gms | Countries can adapt the definition according to local requirements |
| 6 | India - Ministry of Health and Family Welfare (MoHFW) | ≥28 weeks |  | Earlier deaths before 28 weeks are classified as miscarriages |

**Supplementary Table 2:**Details of principal investigators contributing data to the ICMR-SPIC dataset

| **Sr. No** | **Cohort name** | **Site / Institute** | **Name of PI and Co-PIs** |
| --- | --- | --- | --- |
| 1 | CalPreg | St. Johns’ Research Institute (SJRI),Bengaluru | Dr Pratibha Dwarkanath  Dr AnuraV Kurpad |
| 2 | GARBHINI | Translational Health Science and Technology Institute (THSTI), Faridabad | Dr Shinjini Bhatnagar  Dr Nitya Wadhwa  DrRamachandran Thiruvengadam  (PIMS, Puducherry) |
| 3 | LIFE | MediCiti Institute of Medical Sciences, Ghanpur, Telangana | Dr KalpanaBasany |
| 4 | MAASTHI | Indian Institute of Public Health (IIPH)-Bengaluru, Public Health Foundation of India | Dr Giridhar R Babu  Dr Debarati Mukherjee |
| 5 | MaatHRI | University of Oxford, UK | Dr Manisha Nair |
| 6 | MNHR-Belagavi | Jawaharlal Nehru Medical College (JNMC), Belagavi; KLE Academy of Higher Education and Research, Belagavi | Dr Shivaprasad S Goudar  Dr ManjunathSomannavar  Dr Sangappa M Dhaded  Dr Avinash Kavi |
| 7 | MNHR-Nagpur | Lata Medical Research Foundation, Nagpur | Dr Archana Patel  Dr Kunal Kurhe |
| 8 | PMNS | King Edward Memorial (KEM) Hospital and Research Centre, Pune | Dr Chittaranjan S Yajnik  Dr Urmila Deshmukh |
| 9 | REVAMP | Interactive Research School for Health Affairs (IRSHA), BharatiVidyapeeth University, Pune | Dr Sadhana Joshi  Dr Girija Wagh  Dr Sanjay Lalwani  Dr Sanjay Gupte  Dr Juhi Nema |
| 10 | WINGS | Society for Applied Studies, New Delhi | Dr Nita Bhandari  Dr Ranadip Chowdhury |
